# Aortic valve stenosis induced occult hemoglobin release promotes endothelial dysfunction

**DOI:** 10.1101/2022.12.01.22282891

**Authors:** Christine Quast, Florian Bönner, Amin Polzin, Verena Veulemans, Isabella Gyamfi Poku, Ramesh Chennupati, Magdalena Nankinova, Nicole Staub, Juliane Jokiel, Fabian Keyser, Jasmina Hoffe, Katrin Becker, Pia Leuders, Saif Zako, Ralf Erkens, Christian Jung, Ulrich Flögel, Michael Neidlin, Ulrich Steinseifer, Sven Thomas Niepmann, Sebastian Zimmer, Martin Feelisch, Tobias Zeus, Malte Kelm

## Abstract

**Rationale:** The impact of aortic valve stenosis (AS) on systemic endothelial function independent of standard modifiable risk factors (SMuRFs) is unknown.

**Objective:** We hypothesized that AS induces subclinical hemoglobin release from red blood cells (RBCs) following transvalvular passage due to post-stenotic aberrant blood flow and that cell-free hemoglobin (fHb) may limit endothelial NO bioavailability, affecting vascular function.

**Methods and Results:** AS induces swirling blood flow in the ascending aorta which impairs RBC integrity with consecutive release of fHb. Indeed, swirl flow magnitude assessed by 4D flow cardiac magnetic resonance correlates with fHb levels. Elevated systemic fHb reduces NO bioavailability and thus impairs endothelial cell function as evidenced by impaired flow mediated dilation (FMD). In addition, we here demonstrate impaired FMD in an experimental model of AS utilising C57BL/6 mice with preserved left ventricular function and without cardiovascular risk factors. In this model, endothelial dysfunction is accompanied by significantly increased fHb, exaggerated NO consumption and increased plasma levels of nitroso species and the final NO oxidation product, nitrate. Scavenging of fHb by infusion of haptoglobin reversed these deleterious effects. There observations were verified by transfer experiments with human plasma (sampled from patients with AS sheduled for TAVR) using a murine aortic ring bioassay system where the plasma from AS patients induced endothelial dysfunction when compared to plasma from control individuals without AS. Importantly, these deleterious effects were reversed by successful aortic valve replacement via TAVR independent of SMuRFs.

**Conclusions:** In aortic valve stenosis, increases in post-valvular swirl blood flow in the ascending aorta induces subclinical hemolysis that impairs NO bioavailability. Thus, AS itself promotes systemic endothelial dysfunction independent of other established risk factors. Transcatheter aortic valve replacement limits NO scavenging by realigning of postvalvular blood flow to normal physiological patterns.

## Introduction

While AS is frequently associated with cardiovascular risk factors such as arterial hypertension (HT), hyperlipoproteinemia (HLP) and diabetes mellitus (DM) – potentially causing endothelial dysfunction – the impact of AS itself on systemic endothelial function remains unknown. AS is characterized by supravalvular turbulences and swirling blood flow in the ascending aorta due to narrowing of the valve orifice, impacting RBC membrane integrity. Disruption of red blood cell (RBC) integrity due to enhanced shear stress can lead to loss of function and release of hemoglobin (fHb) into the extracellular space (1). Cell-free hemoglobin (fHb) and Hb containing microparticles act as nitric oxide (NO) scavengers (2), and therefore affect the bioavailability of NO, with unfavorable consequences for vascular and endothelial homeostasis (3). However, blood flow simulations by Vahidkhah et al. suggested that in AS hemolysis is unlikely and that the shear-stress induced damage to RBCs is mostly below the hemolytic threshold (4). In contrast, Meyer et al. evaluated the impact of blood flow on NO bioavailability and demonstrated that flow alterations during hemodialysis induce the release of hemoglobin with consecutive endothelial dysfunction due to disturbed NO bioavailability (5).

Using a combination of animal experimentation, observations in patients and human biospecimen/animal tissue bioassay transfer experiments the present study aimed to 1) exclude or identify endothelial dysfunction in experimental AS using a well defined murine model of aortic valve stenosis free of confounding cardiovascular comorbidities, 2) reveal whether AS is associated with a dysregulation of the systemic NO pool and whether or not this is related to blood flow heterogeneity, and finally 3) test the hypothesis that these alterations can be reversed by application of haptoglobin and/or valve replacement.

## Methods

### Animals

Male 12 week-old C57Bl/6 mice (20-28 g body weight) were used in all experiments. Animal experiments were performed in accordance with the national guidelines on animal care and were approved by the Landesamt für Natur, Umwelt und Verbraucherschutz (LANUV, Nordrhein-Westfalen, Germany) under file reference 84-02.04.2017.A172. All animals used in this study were purchased from Janvier, used after a minimum of 7 days oc acclimatization to local vivarium conditions and kept at a normal 12/12h light/dark cycle at the central animal research facility of the Heinrich-Heine University, Düsseldorf, Germany. They were fed with a standard rodent chow and received tap water *ad libitum*. Numbers per groups are as indicated in the results section and figures.

### Patient and control cohort

1.554 Patients gave their informed consent according to the Declaration of Helsinki and the German Aortic Valve Registry (GARY). Ethics committee of Heinrich-Heine University Duesseldorf, Germany, gave ethical approval for this work (No. 2018-86 // ID: 2017114515, No. 5761R, ID 2016105862). All patients gave their informed consent. CT was performed prior to TAVR according to routine diagnostic workflows. In a subcohort, 305 patients underwent TAVR, of whom 293 participated in further analyses. Cardiovascular Magnetic Resonance (CMR) analysis was performed in n=25 patients with AS before TAVR and n=18 patients after TAVR. 30 age- and comorbidity-matched patients were enrolled in the control group and gave their informed consent. Patient characteristics of the patients who entered the final multivariate analysis are shown in Table 1.

**Table 1.**
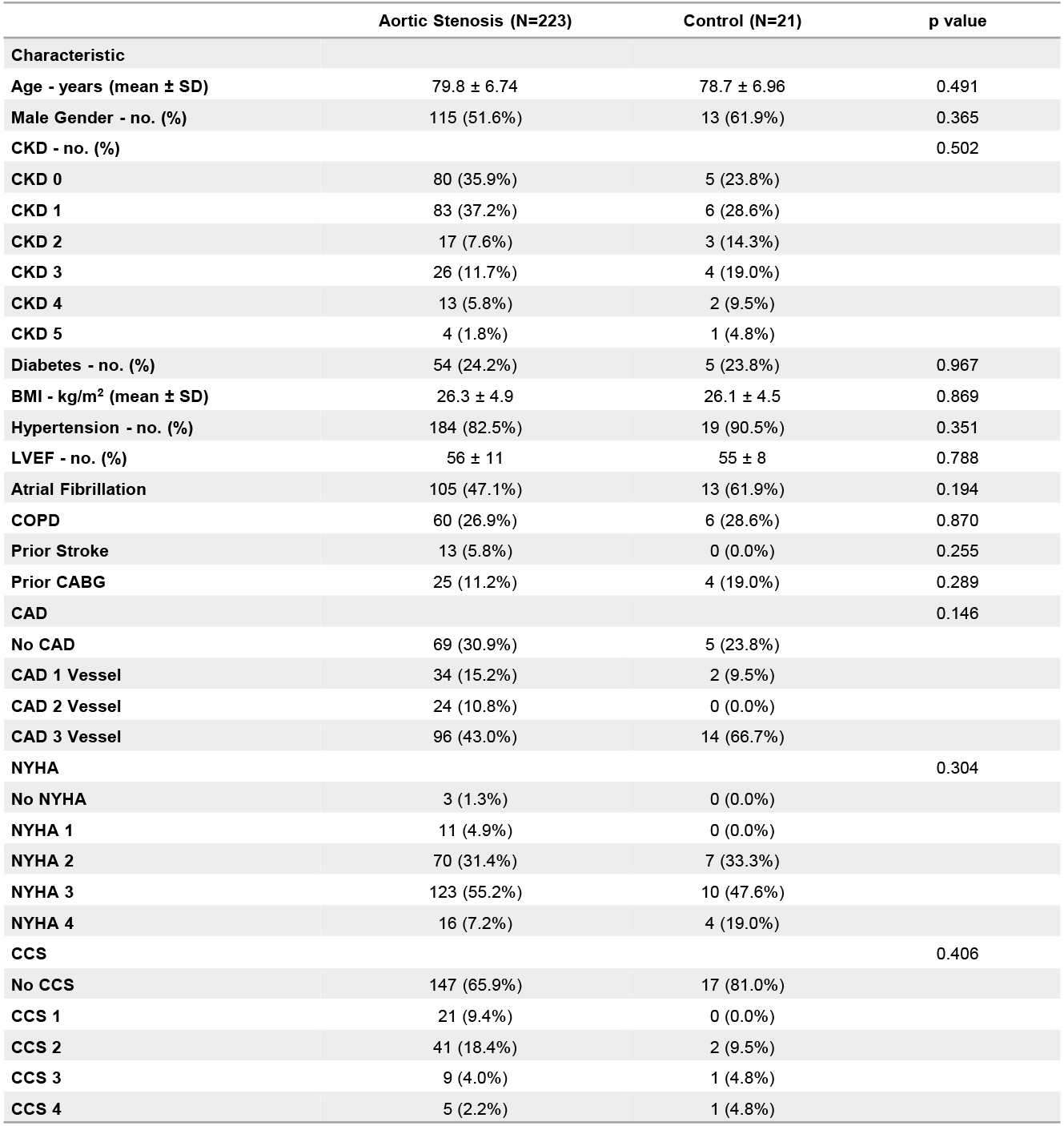
Patient characteristics. The table shows the patient characteristics of the cohort included in the final multivariate analysis. The number of patients with AS (before and after TAVR) was n=223 whereas n=21 subjects without AS were included in the age- and comorbidity-matched control cohort.

|  | Aortic Stenosis (N=223) | Control (N=21) | p value |
| --- | --- | --- | --- |
| Characteristic |  |  |  |
| Age - years (mean ± SD) | 79.8 ± 6.74 | 78.7 ± 6.96 | 0.491 |
| Male Gender - no. (%) | 115 (51.6%) | 13 (61.9%) | 0.365 |
| CKD - no. (%) |  |  | 0.502 |
| CKD 0 | 80 (35.9%) | 5 (23.8%) |  |
| CKD 1 | 83 (37.2%) | 6 (28.6%) |  |
| CKD 2 | 17 (7.6%) | 3 (14.3%) |  |
| CKD 3 | 26 (11.7%) | 4 (19.0%) |  |
| CKD 4 | 13 (5.8%) | 2 (9.5%) |  |
| CKD 5 | 4 (1.8%) | 1 (4.8%) |  |
| Diabetes - no. (%) | 54 (24.2%) | 5 (23.8%) | 0.967 |
| BMI - kg/m<sup>2</sup> (mean ± SD) | 26.3 ± 4.9 | 26.1 ± 4.5 | 0.869 |
| Hypertension - no. (%) | 184 (82.5%) | 19 (90.5%) | 0.351 |
| LVEF - no. (%) | 56 ± 11 | 55 ± 8 | 0.788 |
| Atrial Fibrillation | 105 (47.1%) | 13 (61.9%) | 0.194 |
| COPD | 60 (26.9%) | 6 (28.6%) | 0.870 |
| Prior Stroke | 13 (5.8%) | 0 (0.0%) | 0.255 |
| Prior CABG | 25 (11.2%) | 4 (19.0%) | 0.289 |
| CAD |  |  | 0.146 |
| No CAD | 69 (30.9%) | 5 (23.8%) |  |
| CAD 1 Vessel | 34 (15.2%) | 2 (9.5%) |  |
| CAD 2 Vessel | 24 (10.8%) | 0 (0.0%) |  |
| CAD 3 Vessel | 96 (43.0%) | 14 (66.7%) |  |
| NYHA |  |  | 0.304 |
| No NYHA | 3 (1.3%) | 0 (0.0%) |  |
| NYHA 1 | 11 (4.9%) | 0 (0.0%) |  |
| NYHA 2 | 70 (31.4%) | 7 (33.3%) |  |
| NYHA 3 | 123 (55.2%) | 10 (47.6%) |  |
| NYHA 4 | 16 (7.2%) | 4 (19.0%) |  |
| CCS |  |  | 0.406 |
| No CCS | 147 (65.9%) | 17 (81.0%) |  |
| CCS 1 | 21 (9.4%) | 0 (0.0%) |  |
| CCS 2 | 41 (18.4%) | 2 (9.5%) |  |
| CCS 3 | 9 (4.0%) | 1 (4.8%) |  |
| CCS 4 | 5 (2.2%) | 1 (4.8%) |  |

### Induction of experimental AS in mice

Mice were anesthetized by intraperitoneal injection of ketamine (100mg/kg) and xylazine (10mg/kg). After endotracheal intubation and mechanical ventilation at a rate of 150 to 180 strokes/min and a tidal volume ranging from 220 to 280 μL, animals were placed in a supine position on a warming pad. Anesthesia was maintained by 1.5 to 2.0 vol% isoflurane. Following skin incision and blunt preparation, the right carotid artery was exposed under an upright dissecting microscope, a coronary wire was inserted and after passage past the aortic valve rotated under echocardiographic monitoring to induce an aortic valve stenosis as previously described (6, 7). Thereafter, the carotid artery was ligated. Mice were compared to age-matched animals subjected to the same procedure except for passage of the valve (sham) or without surgery (control).

### Functional characterization of mice by echocardiography, flow mediated dilation and invasive blood pressure analysis

Echocardiography was performed before, at one and four weeks after wire injury or sham surgery, respectively. Valvular, myocardial and systemic hemodynamic characteristics were assessed in B-mode, M-mode, colour and pulsed wave doppler under anesthesia with 2% isoflurane administered through a face mask. Successful induction of aortic valve stenosis was confirmed by a significant increase in peak flow velocity as well as peak and mean pressure gradient across the valve as previously described (7). Mice with more than mild aortic regurgitation were excluded from further analysis.

Flow mediated dilation (FMD) in mice was performed in the left femoral artery before and four weeks after induction of the aortic valve stenosis or sham surgery, respectively. Occlusion of the left hindlimb was assured by inflating the cuff above systolic blood pressure (250 mmHg). After five minutes of occlusion the cuff was released. Vessel diameter and flow through the femoral artery were measured every 30 seconds for five minutes before and after cuff release by ultrasound proximal to the cuff. For regain-of-function experiments haptoglobin (phenotype 1-1, 40 μM/kg body weight, Sigma-Aldrich, USA) or vehicle was administered intravenously one minute prior to cuff release.

For invasive blood pressure analysis animals were anesthetized by subcutaneous injection of buprenorphine (0.1 mg/kg body weight) and isoflurane inhalation via a ventilator (micro ventilator, UNO, the Netherlands) with 2 % isoflurane (v/v) with oxygen-enriched room air. The common carotid artery was exposed by blunt dissection followed by temporary ligation of the vessel. After incision of the vessel, the catheter (SPR-829 Mikro-Tip® Millar, USA) was carefully inserted into the vessel. The pressure was recorded for five minutes using the Lab Chart software. Mean arterial pressure was calculated as MAP=2xEDP-ESP/3. Systemic vascular resistance was estimated by the simplified equation SVR≈MAP/CO. Blood pressure was recorded for five minutes (Lab Chart 7, A.D. Instruments, New Zealand).

### Ex vivo characterization of myocardial contractility

Hearts were excised, dissected in prechilled and oxygenated buffer and connected to a Langendorff apparatus (Hugo Sachs Electronics, March-Hugstetten, Germany) by cannulation of the ascending aorta as previously described (8). Retrograde perfusion of the isolated hearts with modified Krebs–Henseleit buffer was immediately started. An electrode was fixed for pacing the hearts at a constant frequency of 600 min. After an equilibration period hearts were perfused with isoproterenol with a final concentration of 12 nmol according to standardized protocols. Pressure was transmitted by a non compaction balloon placed in the left ventricle.

### Blood collection, FACS, cell-free hemoglobin and haptoglobin

#### Mice

Blood was taken by retroorbital puncture for blood count or by cardiac puncture for FACS, NO consumption and hemoglobin measurements four weeks after wire injury or sham surgery, respectively. Samples with iatrogenic hemolysis were excluded. Cell-free hemoglobin and haptoglobin was analyzed in plasma with a commercially available enzyme-linked immunosorbent assay kit (abcam) according to the manufacturer’s instructions. Red blood cells were washed and analyzed using FACS to detect externalization of phosphatidylserine by binding of annexin V (Annexin V-PE, BD Bioscience). Furthermore, to distinguish “old” (CD71 positive) from “young” (CD71 negative) RBCs expression of cell surface marker CD71 (CD71-APC, MACS Miltenyi Biotec) was used. Plasma aliquots used for measurement of NO consumption were stored at -80° before analysis.

#### Humans

Venous blood was collected before and 1-3 days after TAVR. For regional fHb level analysis, blood was taken by a cardiac catheter situated in the left ventricle (LV), immediately above the valvular cusps (Valve), the ascending (Ao asc) and descending aorta (Ao desc), respectively, before and after TAVR. Analysis of cell-free hemoglobin and haptoglobin was performed by the local Central Institute for Clinical Chemistry and Laboratory Diagnostics, University Hospital Düsseldorf. FACS analysis was performed using RBC specific markers such as CD235a (APC, Biolegend), Annexin-V (PE-Cy7, Biolegend) and CD71 (FITC, Miltenyi Biotec) according to standardized protocols. For transfer experiments, plasma was separated from RBCs by centrifugation for 10 minutes at 800 xg and 4 °C. RBCs were washed twice with PBS.

### NO consumption assay

The extent to which plasma consumes nitric oxide (NO) was assessed using an NO consumption assay, essentially as described (9) except that we used a different chemiluminescence detector (CLD 88, Eco Medics). Briefly, under deep isoflurane anesthesia (3.5%), blood (1 ml) was collected in heparin-coated tubes from mice with and without aortic stenosis. The blood was centrifuged at 800 xg for 10 min at 4 °C, the buffy coat was removed, and plasma and RBC pellet were collected for the NO consumption assay. First, the system was calibrated by injection of known amounts of nitrite into a reduction solution consisting of 45 mmol/L potassium iodide (KI) and 10 mmol/L iodine (I_2_) in glacial acetic acid kept in a septum-sealed reaction chamber at a constant temperature (60 °C) (10, 11); the chamber was continuously bubbled with helium gas. Afterwards, the reaction chamber was cleaned with acetone and PBS. Freshly prepared 120 μM DetaNONOate (Cayman Chemicals) in PBS (pH 7.4) was added to the reaction chamber and continuously purged with helium. After achieving a stable signal for NO release from the NO-donor, plasma (50 μl) samples were injected into the reaction chamber and a transient decrease in baseline NO signal was recorded (indicative of NO consumption). Relative NO consumption levels were calculated based on (negative) areas under the curve and compared to the (positive) values for NO release from known amounts of nitrite standards.

### Human transfer bioassay experiments

Aorta was dissected from healthy 12 week-old C57Bl/6 mice, cut into rings and co-incubated with either plasma or washed red blood cells, with or without haptoglobin (final concentration 5nM/ml solution), from patients with AS before and one to three days after TAVR at 37 °C. RBCs suspension for co-incubation was adjusted to a hematocrit of 40% in modified Krebs Henseleit buffer. Hereafter, aortic rings were carefully rinsed and loaded to the aortic ring apparatus. After equilibration, the vascular tissue was exposed to acetylcholine (ACH, 0.1 nM - 10 μM), phenylephrine (PHE, 0.1 nM – 10 μM) and sodium nitroprusside (SNP, 0.01 nM – 10 μM) according to standardized protocols. All experiments were carried out in the presence of indomethacine (10 mM) to exclude potential prostaglandin-mediated vasoconstrictor effects counteracting NO mediated dilation. Data were recorded by LabChart 8 software.

### CMR Data Acquisition and Analysis

Healthy age-matched volunteer and patient scans were performed using a 1.5 T scanner (Archieva, Philips, Best Netherlands) with a 32-channel phased array coil including four-dimensional (4D) flow acquisition. For analysis, a dedicated software (Circle CVI 42, Circle Cardiovascular Imaging Inc., Calgary, AB, Canada) was used for full automatic delineation of cardiac parameters including 4D flow maps. Within CMR the following parameters were assessed: aortic valve area (AVA, [cm^2^]), maximum wall shear stress in the ascending aorta (WSS, [Pa]), peak velocity in the ascending aorta [cm/s] and viscous energy loss [J/m^3^] calculated from the 3D velocity field with full coverage of the thoracic aorta with region of interest in the ascending aorta (12). Turbulences or swirling flow, respectively, were visualized in 4D flow maps and graded as follows: 0 no turbulence, grade 1 is swirling flow comprising less than 360° of the aortic circumference (= Helix I°), grade 2 is swirling flow exceeding more than 360° of the aortic circumference in the ascending aorta (= Helix II°). Flow displacement was calculated as previously described (13).

### 3D image analysis by multi-slice computed tomography and calcium burden

Multi-slice computed tomography (MSCT) was routinely performed in all patients sheduled for TAVR. CT data were obtained using a 128-slice, single source CT scanner with temporal resolution of 150 ms and a collimation of 128 × 0.6 mm (“SOMATOM Definition AS+”, Siemens Healthcare, Forchheim, Germany) according to TAVR-related standardized recommendations for CT image acquisition (14). Images were analyzed in the diastolic phase and transferred to a dedicated workstation (3mensio Structural Heart™, Pie Medical Imaging BV, Maastricht, The Netherlands). All MSCT-reconstructions and related analyses were performed by experienced level-3 readers. The total AVC and calcium amount of the upper LVOT is expressed as recalculated Agatston units (AU) adapted from the calcium volume. Every area section was handled separately (LVOT, AVC, leaflets) concerning the calcium amount and according to current recommendations. A threshold of at least 500 mm^3^ was set to account for the correlation of fHb with global calcification of the aortic valve, separated into graded calcification (AVC 0 = no calcification until AVC 3 >2.065 AU in men or >1.274 AU in women, respectively, according to (15)).

### Statistics

All data are illustrated as mean values ± standard deviation (SD) or standard error of the mean (SEM) as indicated. To determine significant differences between groups, Student’s t test, t-test according to Wilcoxon, two-way ANOVA, Sidak’s or Tukey’s multiple comparison test, respectively, Mann-Whitney-Test, Kolmogorov-Smirnov-Test analysis, Pearson correlation coefficient and multivariate analysis have been applied as required. P values of ≤0.05 have been considered being statistically significant.

## Results

Comprehensive characterization of an experimental aortic valve stenosis and postvalvular aortic blood flow in AS revealed the existence of 1) endothelial dysfunction in AS secondary to 2) impaired RBCs integrity and increased fHb, which 3) in turn affects the circulating NO pool, 4) is associated with causative increased swirling blood flow in the ascending aorta; finally, 5) endothelial dysfunction readily reversed by treatment options such as valve replacement (TAVR) or fHb scavenging.

### Experimental AS was characterised by endothelial dysfunction in the absence of cardiovascular comorbidities and cardiac functional deterioration

FMD was significantly impaired in experimental AS, indicative of endothelial dysfunction. Specifically, sham mice without AS exhibited physiological FMD response, whereas maximum FMD in AS mice was significantly decreased (Fig. 1A). The NOS inhibitor, L-NAME served as positive control and significantly inhibited FMD in both AS and sham animals, as expected. FMD was not affected by alterations in systemic blood pressure since the latter remained unchanged four weeks after induction of experimental aortic valve stenosis (Figure 1B). Heart rate and systemic vascular resistance (SVR) did not reveal significant differences between sham and mice with AS either (Fig. 1C, D). In addition to these systemic hemodynamic parameters, experimental aortic valve stenosis was characterized by preserved cardiac functional parameters. AS was verified by acquisition of peak velocity as well as peak and mean pressure gradient across the aortic valve (Fig. 1E). While ejection fraction (EF) and left ventricular inner diameter in diastole (LVIDd) were preserved in AS, mice with AS exhibited a mild but significant increase of the interventricular septum in diastole (IVSd) indicating compensatory mild concentric hypertrophy (Figure 1F). When hearts were tested *ex vivo* using a classical Langendorff apparatus, left ventricular contractility (LVDP, dP/dtmax, dP/dtmin) at rest and contractile reserve upon stimulation with isoproterenol were both significantly enhanced in AS (Figure 1G). Taken together, these results demonstrate that induction of experimental AS leads to subtle myocardial adaption to maintain systemic circulatory hemodynamics without overt functional deterioration; yet, these adaptions are associated with markes vascular endothelial dysfunction.

**Figure 1.**
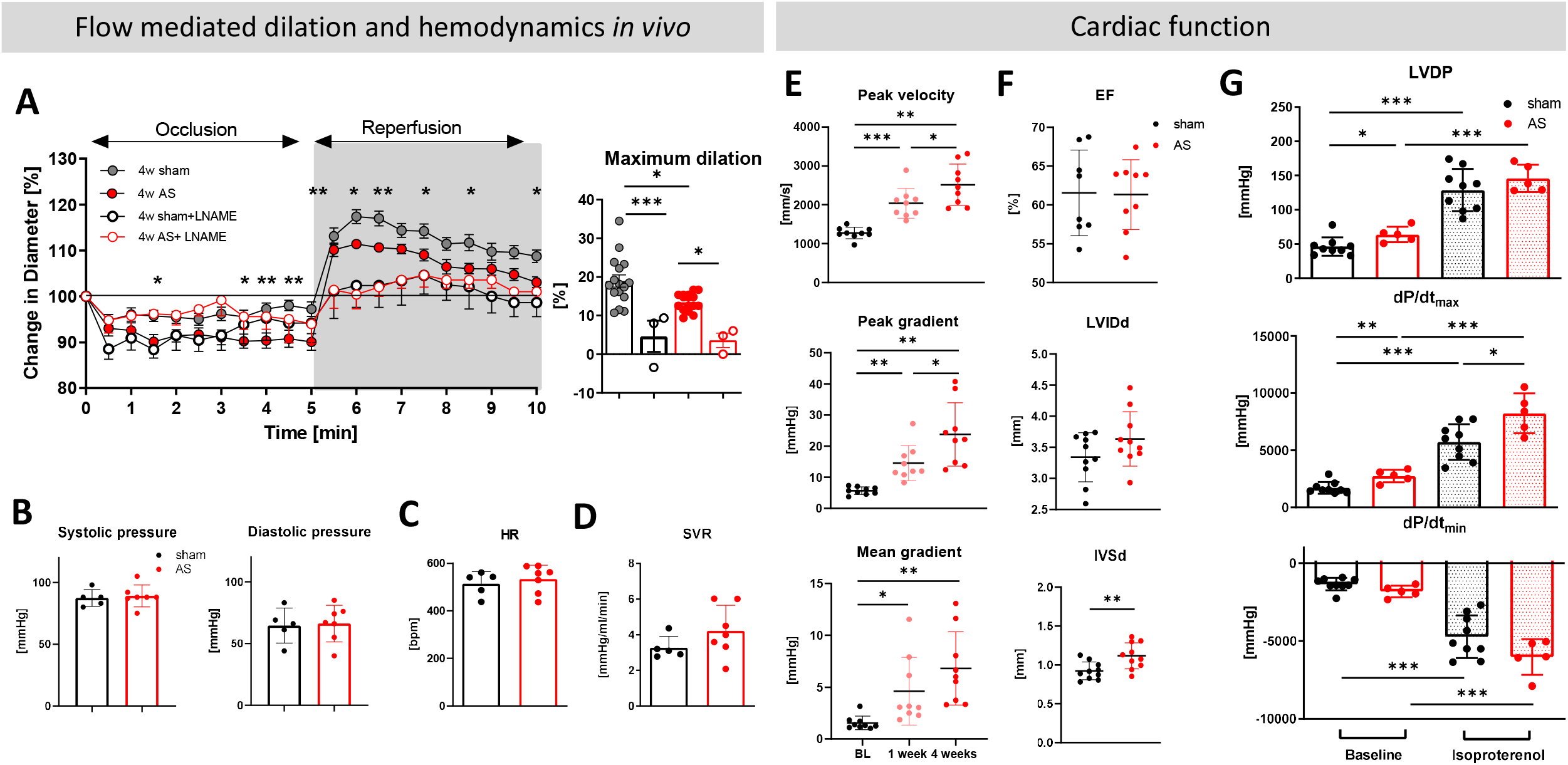
Endothelial dysfunction in experimental AS. A: Flow mediated dilation *in vivo*. Four weeks (4w) after wire injury or surgery without induction of AS (sham) flow mediated dilation of the femoral artery was investigated (sham and AS n=16 each group, n=3 each group with L-NAME, 2-way ANOVA, * sham vs AS, * p<0.05, ** p<0.01). B-D: Systolic and diastolic blood pressure, heart rate (HR) and systemic vascular resistance (SVR) remain unchanged four weeks after induction of AS (n=5-7, p = n.s., unpaired student’s t-test). E-G: Cardiac function in experimental aortic valve stenosis. E: Peak velocity, peak and mean gradient across the aortic valve before (black, Baseline, BL), 1 (light red) and 4 weeks (red) were significantly increased after induction of aortic valve stenosis in c57Bl/6 mice, respectively (*p<0.05, **p<0.01, ***p<0.001). F: Analysis of left ventricular function shows normal ejection fraction (EF) and preseved left ventricular inner diameter in distaole (LVIDd). Significantly increased thickness of interventricular septum in diastole (IVSd) indicates concentric hypertrophy due to pressure overload in AS (p = 0.0054). G: Contractile reserve *ex* vivo. Left ventricular contractility *ex vivo* in Langendorff apparatus (LVDP, dP/dtmax, dP/dtmin) is significantly enhanced in AS at rest (LVDP, dP/dtmax) and under stimulation with isoproterenol (dP/dtmax) compared to sham mice without induction of AS.

### RBC integrity is impaired and impacts the circulating NO pool

Reduced RBC integrity in experimental AS went along with enhanced cell-free hemoglobin levels. Mice with experimental AS showed normal total hemoglobin, RBC distribution width and reticulocyte count (Figure 2A, B, E). However, fHb levels (Fig. 2C) and number of eryptotic RBCs, characterized by Annexin V externalisation (Fig. 2F), were significantly increased in AS while haptoglobin was not significantly different in AS (Fig. 2D). Our experiments demonstrated significantly increased NO consumption in AS mice compared to sham operated animals (Fig. 2G,H). Systemic nitrite levels did not differ between groups in experimental AS while nitrate and nitroso species concentrations were significantly higher in AS (Figure 2I-K).

**Figure 2.**
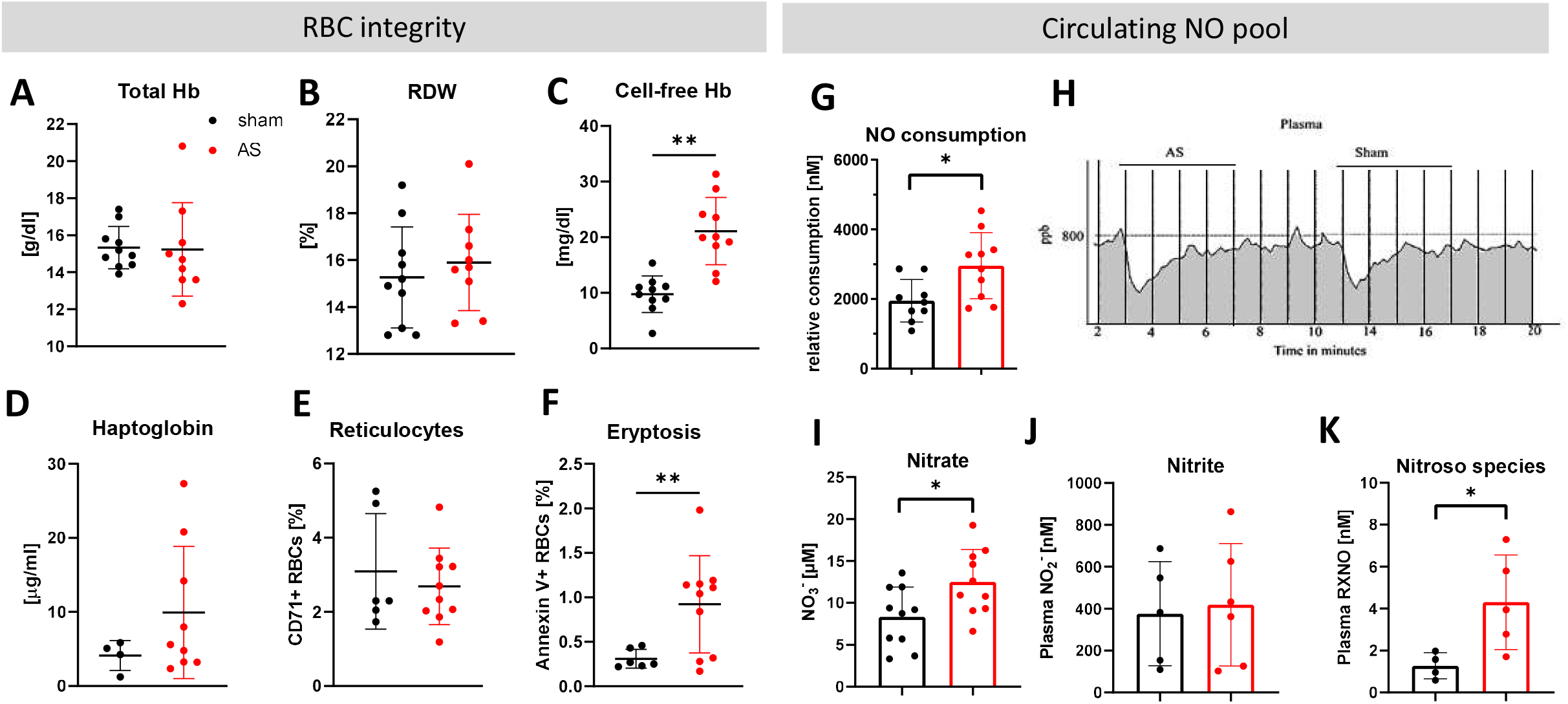
Experimental AS impairs RBCs integrity and impacts circulating NO pool. A-F: RBC integrity, G-K: Circulationg NO pool. A, B: Anemia is not present four weeks after induction of AS as indicated by normal total Hb and RBC distribution width (RDW). C, D: Cell-free hemoglobin levels are significantly increased in experimental AS while haptoglobin remains unchanged. E, F: Reticulocytes (CD71+ RBCs) are not enhanced, while RBC integrity is significantly impaired as shown by increased Annexin V externalisation indicating enhanced programmed cell death (Eryptosis, n=5 and 10, respectively, p<0.05). G-K: Cell-free hemoglobin exaggerates depletion of circulating pool of bioactive NO. G: NO consumption capacity of plasma is significantly increased four weeks after AS induction compared to sham (n=9 sham, n=10 AS, p=0.0145). H: Panels represent original traces during NO consumption assays of AS and sham plasma samples (NO consumption is calculated from AUC, grey). NOx levels show significantly increased levels of nitrate (I) and nitroso species (K) in AS while nitrite (J) is unchanged (mean ± SD, n=5-7, p=n.s.) (values are shown as mean ± SEM, *p≤0.05, unpaired student’s t-test was used to compare between two groups).

Further, *in vivo* experiments demonstratedoft hat impaired endothelial function is reversed upon treatment of animals with haptoglobin, which is able to scavenge free Hb. Reduced flow mediated dilation in mice with AS was fully reversed by administration of haptoglobin just prior to the reperfusion phase (Figure 3) whereas in sham mice, haptoglobin did not show additional FMD anhancement. Application of hemoglobin served as positive control and was comparable to the effects of AS. Application of the vehicle (saline) instead of haptoglobin served as control for potential volume effect showing no significant changes in FMD.

**Figure 3.**
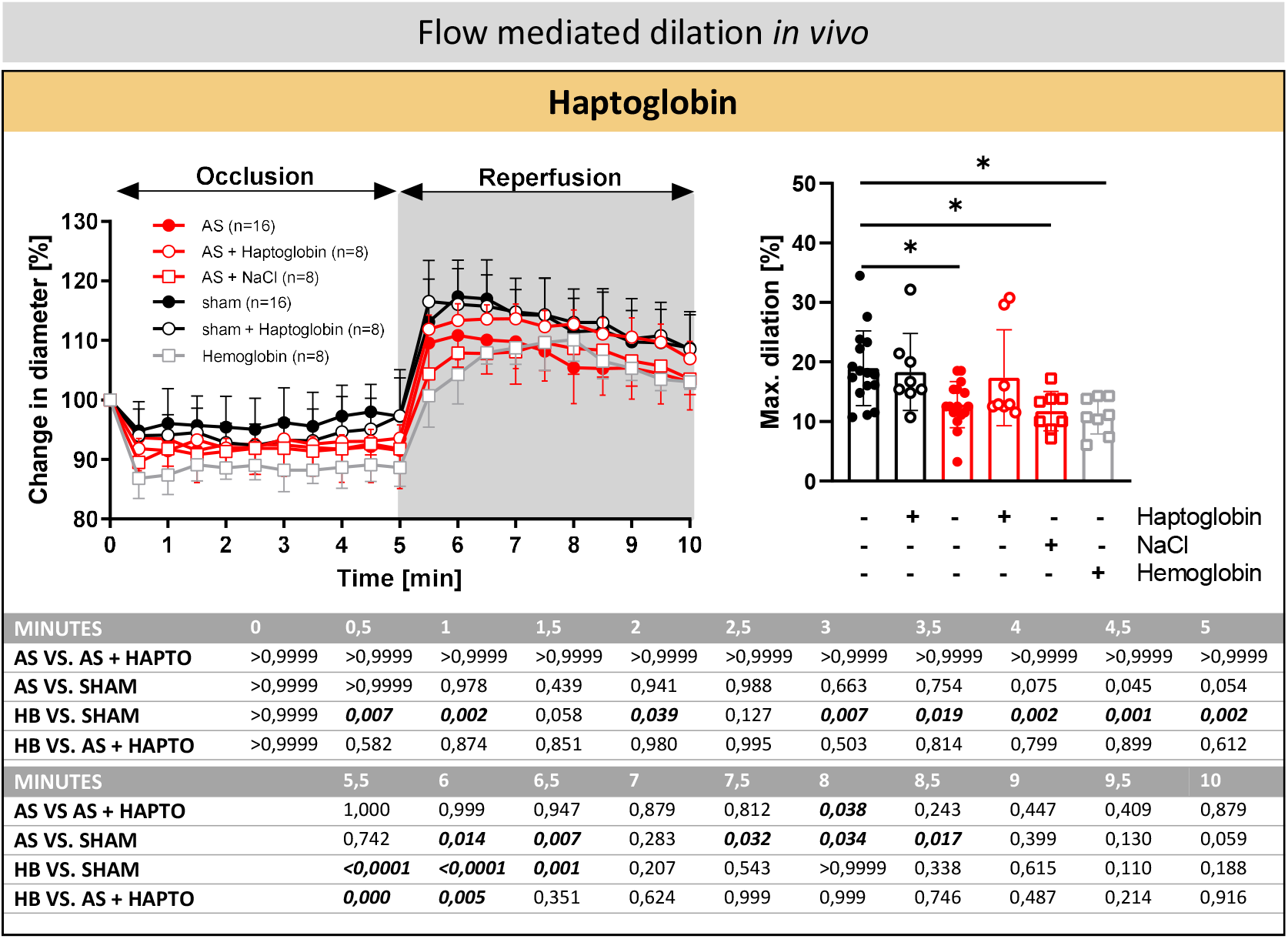
Reversal of endothelial dysfunction in AS by haptoglobin. A: Decreased FMD is partially reversed by the addition of haptoglobin in experimental AS. This effect is not attributtable to volume application as shown by volume control with NaCl in AS. Application of cell-free hemoglobin verifies impaired flow mediated dilation probably due to NO scavenging (p values are depicted in the table below, One-way ANOVA for comparison of maximum dilation [%] between groups).

### Impaired RBC integrity was restored after TAVR

Consistent with our observations in the experimental animal model, circulating fHb levels were found to be significantly higher in patients with AS compared to age/sex-matched controls (Figure 4B). Total Hb count was preserved in severe (>0.6cm^2^) and very severe AS (≤0.6cm^2^) (Figure 4A).

**Figure 4.**
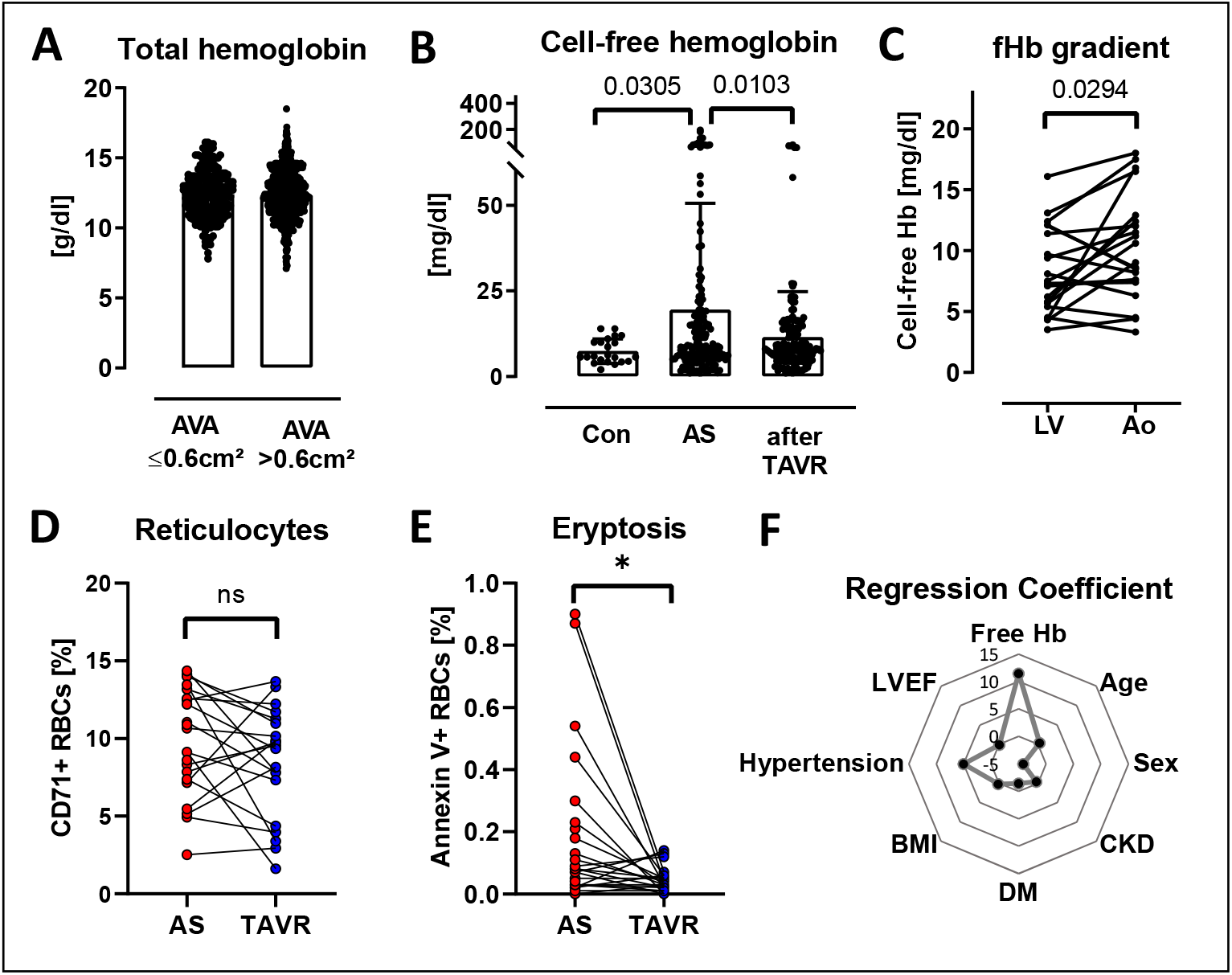
Impaired RBC integrity in AS is restored after TAVR. A: Total hemoglobin is not different between severe (AVA >0.6cm^2^) and very severe AS (AVA ≤0.6cm^2^). B: Cell-free hemoglobin is significantly increased in patients with AS compared to age-matched controls (con) and declines significantly after transcatheter aortic valve replacement (Control n=21, AS n= 158, post TAVR n=155, Kolmogorov-Smirnov-Test). C: Cell-free hemoglobin levels differ between anatomical regions and show a gradient from left ventricle to just above aortic valve (n=21, paired Student’s t-test). D: Fraction of reticulocytes (CD71+ RBCs) is unchanged, but Annexin V externalisation on RBCs indicating eryptosis of RBCs is significantly reduced after TAVR (n=22 before and after TAVR, respectively, paired t-test). F: Regression coefficient analysis of comorbidities in AS reveals strongest regression coefficient for fHb in AS.

Cell-free hemoglobin featured a transvalvular locoregional gradient (Fig. 4C) with fHb levels in patients with AS being significantly increased just above the aortic valve compared to blood sampled from the left ventricle. Again, reticulocytes and ageing RBCs were analysed. Reticulocyte count was the same before and after TAVR (Figure 4D), but in contrast, eryptotic RBC count significantly decreased after TAVR (Figure 4E). Multivariate analysis of comorbidities in AS showed highest regression coefficient for fHb compared to relevant comorbidities (hypertension, adiposity, diabetes mellitus type 2, chronic kidney disease) or patient characteristics such as sex or age (Fig. 4F). The corresponding patient characteristics of the cohort for final multvariate analysis is summarized in table 1.

### Impact of ventricular, valvular and aortic parameters on fHb levels

Our data provided evidence that fHb levels increase with severity of AS from AVA <1.0>0.6 cm^2^ to ≤ 0.6 cm^2^ (Figure 5C). Of note, ejection fraction and calcification of the LVOT did not affect fHb levels (Figure 5F, 5G). Although increasing narrowing of the aortic valve was associated with increased fHb levels, mean pressure gradient across the valve and valvular calcification with high calcium burden did not influence fHb. Yet, aortic flow pattern changes rather dramatically with AS (Figure 5A). In AS, aortic blood flow started to exceed up to or more than 360° of the aortic circumference (grade 2) in the ascending aorta, creating pronounced swirling blood flow. Age-matched healthy are characterized by no helical flow (grade 0) or less than 360° of the aortic circumference (grade 1) (Figure 1D). However, aortic flow displacement, e.g. by eccentric jets, was not associated with increased fHb. Turbulences and swirling flow due to AS were partially reversed after TAVR with less helical flow grade 2 in favour of enhanced grade 1 and 0 (Figure 5B).

**Figure 5.**
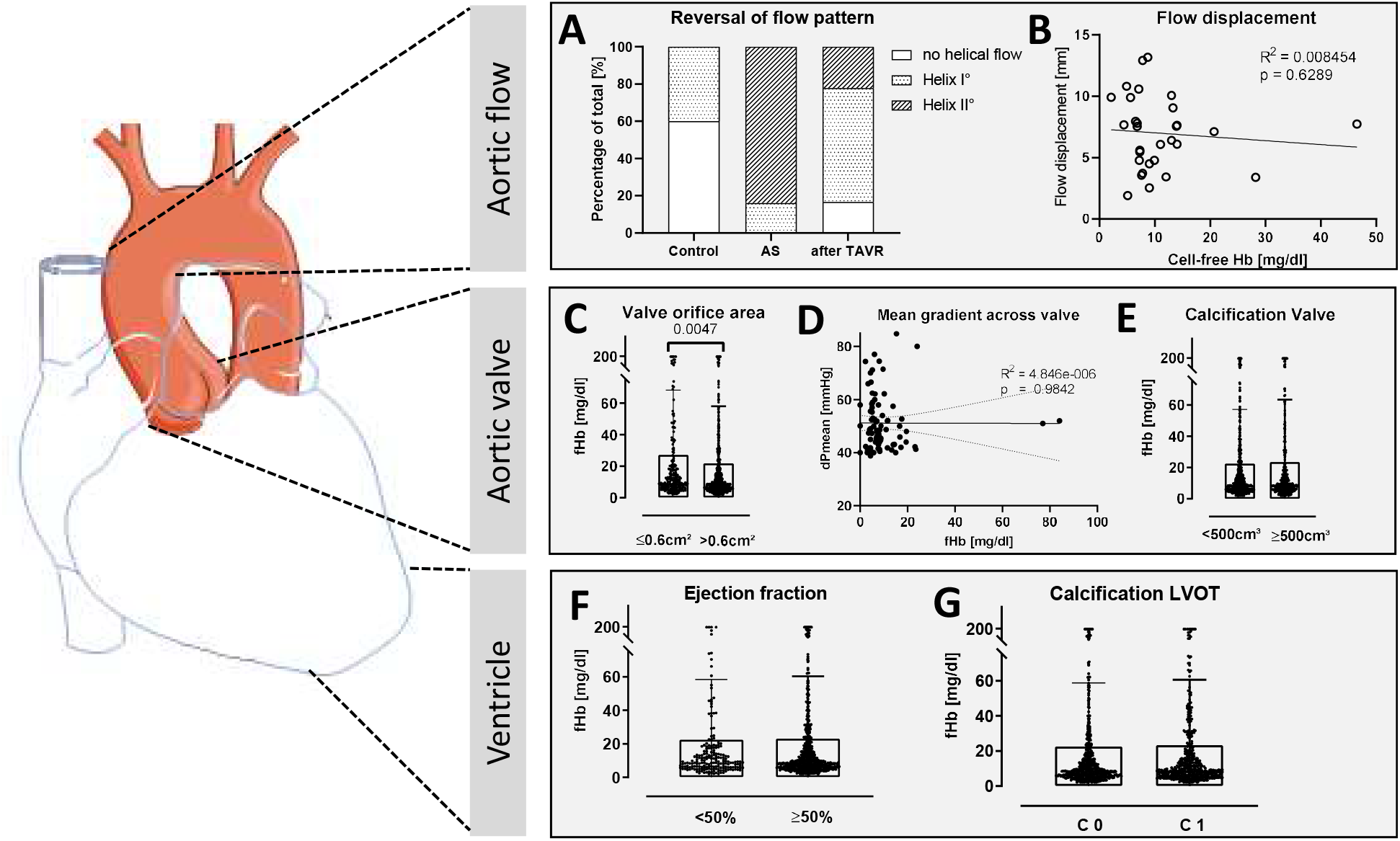
Impact of ventricular, valvular and aortic parameters on cell-free hemoglobin in patients with AS. A: Patients without AS (Control) are characterized by less helical flow in the ascending aorta than patients with severe AS. Helical flow increases in AS and is partially reversed after TAVR (0= no helical flow, Helix grade 1= helical flow <360°, Helix grade 2= helical flow >360°; healthy n=10, AS n=25, TAVR n=18). After TAVR helical flow is less circumferential (Helix I° > Helix II°) than in AS. B: Flow displacement (= deviation of flow from coaxial vessel course) does not correlate with increased fHb. C: fHb level is significantly increased in valves with very severe stenosis (Aortic valve orifice area (AVA) ≤ 0.6cm^2^). D, E: Mean pressure gradient across the aortic valve does not have an impact on fHb levels. fHb levels are independent of a global aortic valve calcium (AVC) burden cut-off of 500 mm2 in patients with severe AS. F, G: Ejection fraction and calcification of left ventricular outflow tract (LVOT) do not increase fHb levels. (mean ± SD, unpaired student’s t-test). (Figure 5 was partly generated using Servier Medical Art, provided by Servier, licensed under a Creative Commons Attribution 3.0 unported license)

### Aortic valve stenosis induces swirling blood flow in the ascending aorta

Patients with aortic valve stenosis exhibit increased forces on the RBCs compared to healthy volunteers without AS due to the higher wall shear stresses and higher peak velocities in the ascending aorta (Figure 6 A,B). The viscous energy loss is increased as well in patients with AS, Figure 6C, which is most probably caused by more pronounced flow disturbances and turbulent flow structures. Moreover, after TAVR flow along the aorta regains homogeneity (Figure 1C).

**Figure 6.**
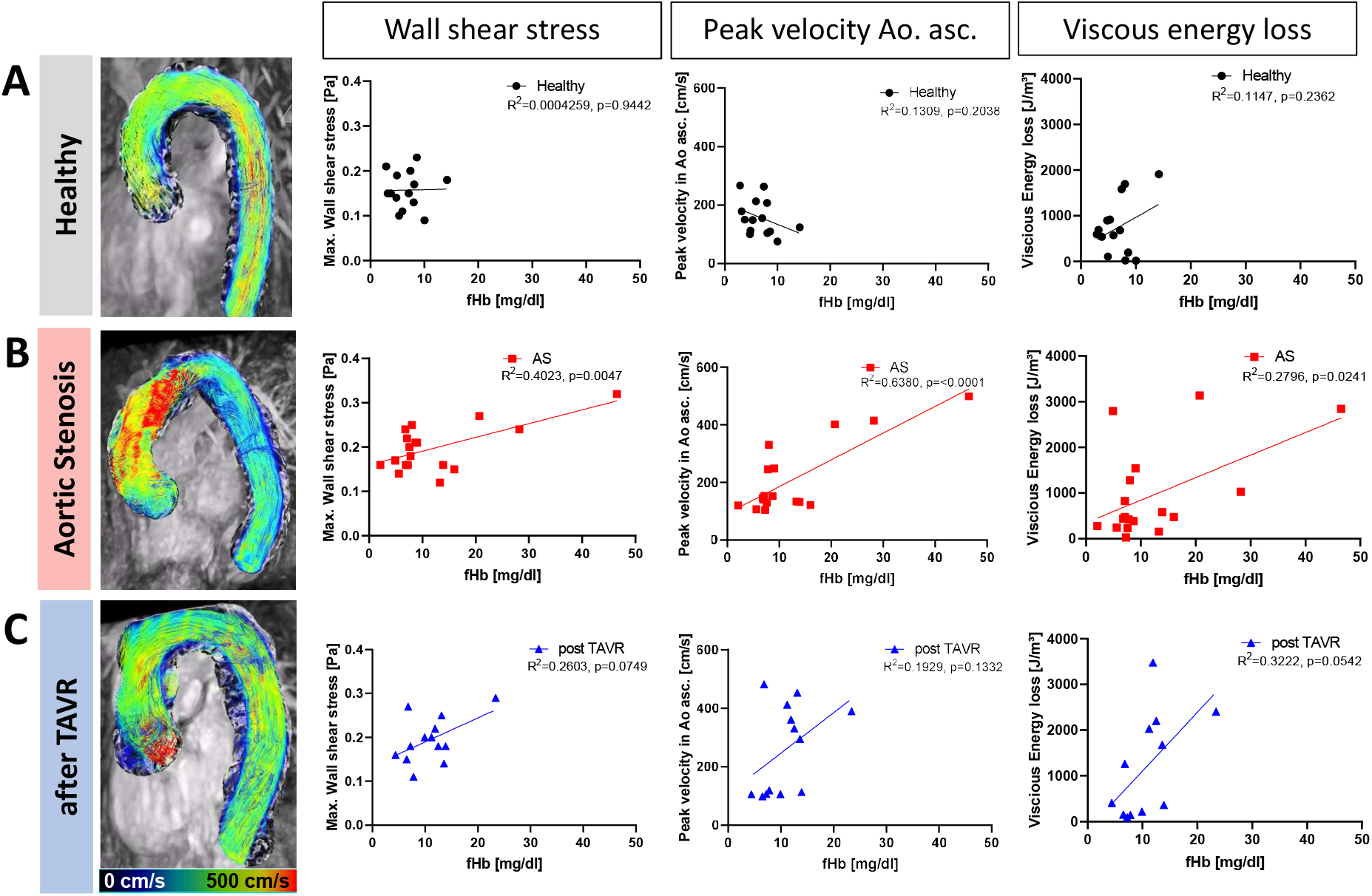
Aortic valve stenosis induces swirling blood flow in the ascending aorta and increases fHb. A: Healthy individuals feature low cell-free Hb levels with physiological wall shear stress, peak velocity in the ascending aorta and viscous energy loss. B, C: Increases in swirling of blood flow in the ascending aorta (post the aortic valve stenosis) indexed by increased wall shear stress, aortic peak velocity and vicious energy loss associated with increasing plasma levels of fHb (WSS: healthy n=14, AS n=18, TAVR n=13; peak velocity in ascending aorta: healthy n=14, AS n=18, TAVR n=13; viscous energy loss: healthy n=14, AS n=18, TAVR n=12, Pearson correlation coefficient).

Healthy individuals feature low fHb levels with physiological wall shear stress, peak velocity in the ascending aorta and viscous energy loss (WSS: CI -0.5156 to 0.5452; peak V Ao. Asc.: CI -0.7486 to 0.2089; Max. viscous energy loss: CI -0.2339 to 0.7369, p=n.s., n=14) (Figure 6A). In AS by constrast, wall shear stress, peak velocity in the ascending aorta and viscous energy loss significantly correlate with fHb levels (WSS: Confidence Interval (CI) 0.2379 to 0.8496, p=0.0047; peak V Ao. Asc.: CI 0.5292 to 0.9219, p<0.0001; Max. viscous energy loss: CI 0.08213 to 0.7985, p=0.0241, n=18) (Figure 6B). Similar tendencies were observed after TAVR (WSS: CI -0.05680 to 0.8283; peak V Ao. Asc.: CI -0.1475 to 0.7973; Max. viscous energy loss: CI -0.009268 to 0.8610, p=n.s., n=13 for WSS and peak velocity in Ao. Asc. and n=12 for max. viscous energy loss) but with less turbulences and lower fHb levels (Figure 6B,C). In summary, healthy patients without exceeding swirling blood flow show physiological fHb levels whereas patients with AS and helical blood flow below or above 360° of the aortic circumference show elevated fHb levels.

### Endothelial dysfunction in transfer experiments improved after treatment options

Transfer experiments with plasma from patients with AS revealed a regain of endothelial vascular reactivity in the bioassay system by haptoglobin (Figure 7A). Similarly, bioassay experiments with RBCs from patients with AS lead to impaired vascular endothelial function compared to RBCs obtained from age-matched controls with similar comorbidities but without AS, and this impairmend was again fully reversed by haptoglobin (Figure 7D).

**Figure 7.**
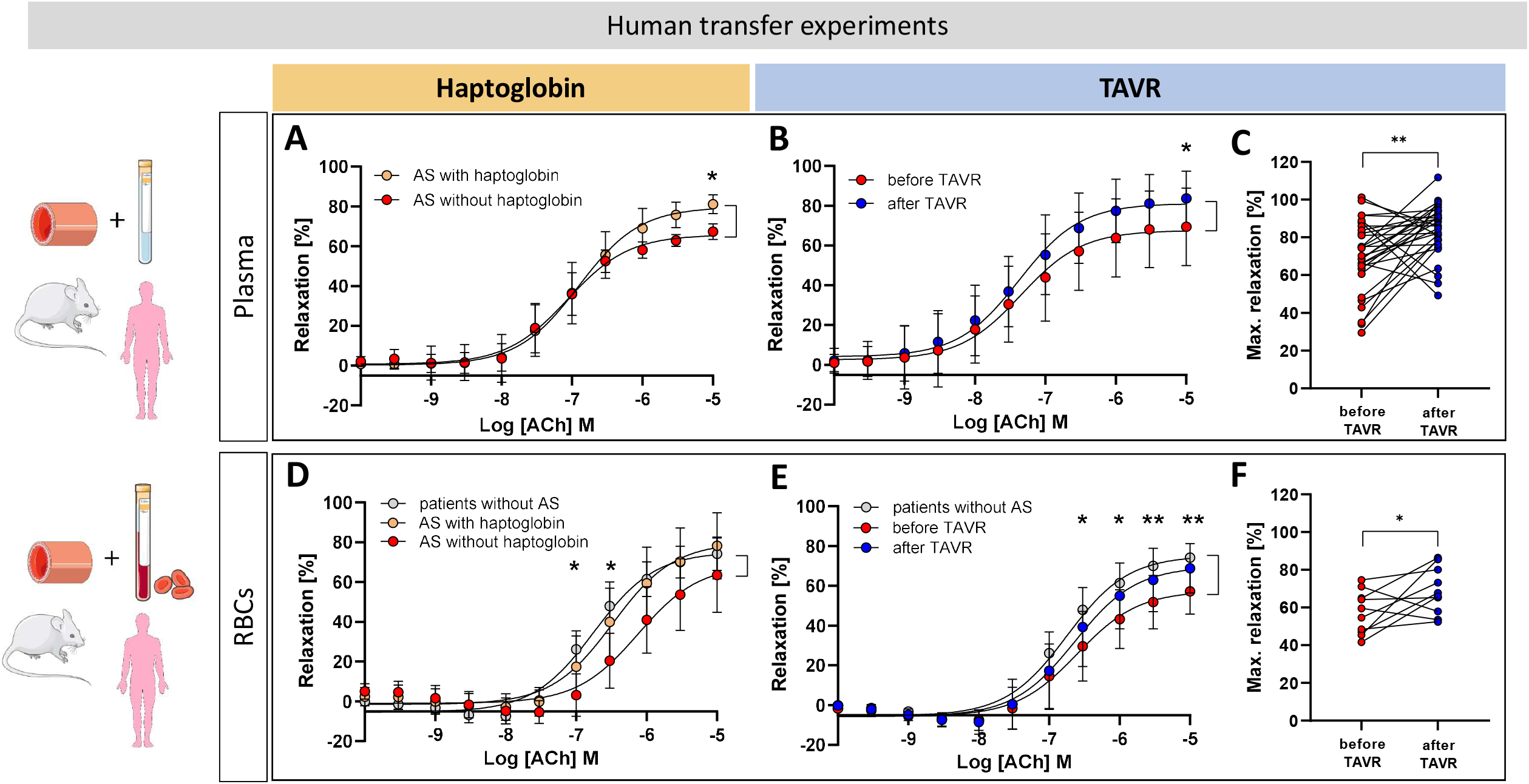
Reversal of endothelial dysfunction in AS. Transfer bioassay experiments with aortic rings from healthy C57Bl/6 mice as target organs are coincubated with plasma from patients with AS (A) or after TAVR (B, C) or RBCs suspension with RBCs from patients with AS (D) or after TAVR (E, F), respectively. A: Co-incubation of plasma with haptoglobin improves endothelial function of aortic rings (n=5 each group, * p<0.05). B: TAVR improves properties of plasma leading to regain of endothelial function showing significantly improved endothelium-dependent relaxation of the target organ in intraindividual comparison (C). D: Coincubation of RBCs from patients with AS with haptoglobin seems to scavenge released fHb since endothelial function regains to the level of patients without AS (patients without AS vs. AS with haptoglobin n.s., * AS without haptoglobin vs. patients without AS, n = 5-7). E: Coincubation with RBCs after TAVR reveals a partial regain of endothelial function compared with AS to a level of RBCs from patients without AS (* patients without AS vs. patients with AS, n = 7-10) in intraindividual analysis (A, B: 2 way ANOVA Sidak’s multipel comparison test; D, E: 2 way ANOVA Tukey’s multiple comparison test; C, F: unpaired student’s t-test).

Moreover, addition of plasma from patients with AS before and after TAVR resulted in significantly improved endothelium-dependent relaxation in the murine bioassay detector system after TAVR in the same patient (Figure 7B, C). We observed similar results with RBCs. Co-incubation of vascular rings with RBCs in the organ bath revealed impaired endothelial function before TAVR. When RBCs were collected from the same patients after TAVR a partial regain of vascular endothelial relaxation was observed, resembling that of RBCs from age-matched controls with comparable comorbidities (Figure 7E, F).

## Discussion

Using a combination of animal experimental results, human observations and transfer bioassay experiments we here demonstrate that 1) experimental aortic valve stenosis is characterized by endothelial dysfunction; 2) swirling blood flow promotes cell-free hemoglobin release in patients with AS; 3) Cell-free hemoglobin due to reduced RBC membrane integrity accounts for reduced systemic NO bioavailability and endothelial dysfunction in AS; and 4) treatment options lead to a marked improvement in endothelial function.

### AS itself induces endothelial dysfunction

It is known that flow disturbances (16) have a critical impact on endothelial function. We are the first to verify that AS itself causes endothelial dysfunction in a model of experimental aortic valve stenosis independent of cardiovascular risk factors, so called SMURFs or relevant comorbidities. To exclude the impact of cardiac functional impairment and potentially confounding comorbidities on endothelial function we used a model of experimental AS without cardiovascular comorbidities and with preserved cardiac and hemodynamic function. Transfer experiments revealed that both blood components, plasma and RBCs, are involved in promoting endothelial dysfunction in AS. While plasma contains increased fHb levels, flow-mediated disturbance of membrane integrity of RBCs due to intensified aortic swirling blood flow may associate with increased fHb release.

### AS compromises RBC integrity

Simulations mimicking abnormal flow due to AS provided information which suggested that the forces acting on blood elements are sufficient to considerably deform red blood cells (17). There is biochemical evidence of subclinical hemolysis in patients with hypertrophic cardiomyopathy, which isassociated with left ventricular outflow tract obstruction provoked by daily physical activities (18), not dissimilar to increased flow velocity in AS. Only a few case reports have evaluated intravascular hemolysis in patients with native valvular heart disease. A study in 30 patients with AS obtained by peak flow velocity reports increased hemolysis without reference to the quality of blood flow, nor a possible reversibility or causality (19).

The increased flow velocity in AS seems to induce mildly disturbed RBC membrane integrity without rupture nor decline of total hemoglobin as detected by phosphatidylserine externalization (Figure 2F, 4E). Maintenance and regulation of asymmetric phospholipid distribution in human erythrocyte membranes are likely important for erythrocyte function (20). In our experiments the disturbed membrane integrity of RBCs may lead to release of cell-free hemoglobin in a manner that stays well below hemolytic thresholds: mice with AS do not display clinically relevant anemia, nor activated erythropoiesis on the basis of normal reticulocyte count, but display significantly elevated f Hb levels (Figure 2 C).

### Pronounced flow disturbances promote hemoglobin release

It is known that patients with aortic valve stenosis exhibit pathological vascular forces in the aorta (21, 22). We showed that pronounced swirling blood flow in the ascending aorta promotes hemoglobin release in patients with AS. Circulating fHb levels increased with higher degrees of swirling blood flow in the ascending aorta assessed by 4D flow MRI. Moreover, we gained evidence for a locoregional gradient of fHb across the aortic valve, with higher levels just above the aortic valve compared to those in the left ventricle. Other flow disturbances like dialyses have been shown to be associated with release of fHb (5). TAVR alters ascending aortic blood flow and wall shear stress patterns (23) and results in less turbulent and helical blood flow (24). In line with this, analysis of calcification (aortic valve, cusps, LVOT) and reduced ejection fraction clearly showed no impact on fHb levels in our study (Figure 5). We hypothesize, that fHb is most likely released by RBCs through either membrane pores or stress-induced membrane blebbing induced by increased helical blood flow above the valve in AS. While fHb release caused by hemolysis is an acknowledged pathomechanism, subhemolytical fHb release is not well understood. Computational simulations demonstrated that shear-induced tensions in AS at the RBC membrane surface are unlikely to cause rupture, but likely to induce membrane plasticity failure (4). Whereas RBC membrane blebbing (with resulting release of intracellular content) is a well-recognized effect of erythrocytic stress imposed by chemotherapeutics/antibiotics (25-27) first models are also arising to calculate fHb release out of transient pores (28).

There is evidence supporting the hypothesis that exposure to supra-physiological shear stress allows for the leakage of RBC intracellular contents (29). But to date, prediction algorithms for complex flows cover pathophysiology incompletely. Specifically, they are unable to predict how different types of fluid stresses translate to red cell membrane failure (30).

### Extracellular hemoglobin accounts for endothelial dysfunction in AS

Free Hb acts as effective NO scavenger nearby the endothelium (1) and might be a potentially crucial mechanism that gives rise to endothelial dysfunction in AS. There are observations that aortic valve stenosis is accompanied by endothelial dysfunction (31). Impaired flow mediated dilation is known as a relevant cardiovascular marker for prognosis – even in emerging infectious disease settings such as the COVID-19 pandemic (32) – and may be associated with severity of AS (33). Recent reports indicate that endothelial dysfunction reverses after interventional valve replacement (31), possibly due to a variety of reasons such as improved blood pressure or forward stroke volume after TAVR. Reduced valvular pressure gradients with less hemodynamic shear stress by TAVR seem to beneficially affect endothelial and vascular homeostasis (34). Similar observations apply to surgical valve replacement in long term analyses (16). But until now, the underlying mechanisms have remained obscure. Comorbidities being persistent in patients with AS may themselves account for endothelial dysfunction (e.g., diabetes mellitus or chronic renal failure (35-37)), and heart failure with reduced ejection fraction might affect endothelial function as well (38). To date, the impact of AS itself on endothelial function had never been considered.

Focussing on fHb as a major cause for endothelial dysfunction in AS, we here unequivocally demonstrate that hemoglobin release into the ascending aorta is triggered in AS, both in experimental aortic valve stenosis (Figure 2C) and in patients with AS (Figure 4B). In line with studies demonstrating fHb to inhibit acetylcholine-induced dilatation of coronary arteries (39), we observed impaired eNOS dependent flow mediated dilation in femoral arteries of mice with AS and healthy mice after application of cell-free hemoglobin verifying fHb as relevant factor for endothelial dysfunction in AS (Figure 3A).

Our observations concerning fHb levels and 4D flow magnetic resonance imaging after TAVR are supported by bioassay analysis with intraindividual comparison of plasma and RBCs in AS (Figure 7 A-F). Once exposed to increased swirling blood flow in AS, RBCs seem to maintain the capacity of enhanced cell-free hemoglobin release as shown by reduced endothelial relaxation in our bioassay experiments with RBCs from patients with AS indicating fHb as key player. On the other hand, plasma from patients with AS shows increased fHb levels and leads to a similar deterioration of endothelial function as being triggered by RBCs.

### Endothelial dysfunction is reversed by treatment options

Finally, our human transfer bioassay experiments revealed a regain of endothelial dysfunction by treatment options by 1) scavenging fHb both in plasma and RBC during co-incubation with haptoglobin or 2) reduced fHb levels due to improved flow conditions following TAVR.

Endothelial dysfunction was reversed in transfer bioassay experiments by co-incubation of rings with plasma and haptoglobin and thereby proving cell-free hemoglobin as relevant origin of endothelial dysfunction in AS. As part of co-incubation with RBCs, addition of haptoglobin resulted in enhanced endothelial function, emphasizing reduced RBC membrane integrity. Haptoglobin might scavenge and thereby counteract hemoglobin which might be released continuously in bioassay, e.g. through pores in disrupted RBS membranes. Haptoglobin acts as an endogenous scavenger of cell-free hemoglobin and protects against fHb mediated endothelial damage (1, 40, 41) and reduced NO bioavailability. In our study, endogenous haptoglobin concentrations were not significantly decreased in AS (Figure 2D) which may be explained by haptoglobin not being relevantly exploited as a counter-measure by elevated cell-free hemoglobin levels in AS. However, in AS, haptoglobin seems to convey local and functionally relevant scavenging of cell-free hemoglobin and thereby enhance local NO bioavailability nearby the endothelium resulting in improved endothelial function. This hypothesis is supported by our *in vivo* experiments with improved FMD in mice with AS after application of haptoglobin prior to cuff release (Figure 3): we demonstrated that intravenous application of haptoglobin in mice with AS and endothelial dysfunction permits a regain of endothelial function. Furthermore, in transfer experiments TAVR resulted in regain of endothelial function as well. Plasma from patients after TAVR exhibited enhanced endothelium relaxation capacity compared to the plasma before TAVR in the same patient (Figure 7B,C). Again, we saw the same result with RBCs from patients after TAVR compared to the same patients before TAVR (Figure 7E,F).

### Limitations of the study

Of note, NO bioavailability can also be affected by complementary mechanisms induced by flow alterations including endothelial microparticles formation (42) owing the capacity to carry heme representing a source of oxidant stress for the endothelium (43). They have been suggested to play a role in AS (31, 34). However, current results on microparticle formation after TAVR is inconsistent with such a mechanism (44). More detailed studies are required to characterize in what form fHb exists in plasma of AS patients.

## Summary and clinical perspective

Taken together, we are the first to demonstrate that endothelial dysfunction is caused by AS itself and is reversed by fHb scavenging indicating enhanced concentrations of extracellular hemoglobin and reduced NO bioavailability as key mechanism underlying endothelial dysfunction in AS; those elevated levels are caused by increased swirling blood flow irrespective of comorbidities.

Cell-free hemoglobin and therefore reduced NO bioavailability appears to be a major determinant of endothelial dysfunction in AS. The documented changes in the circulating NO pool corroborate the functional results and confirm that extracellular hemoglobin impairs NO bioavailability in AS, leading to endothelial dysfunction. The demonstration in transfer bioassay experiments that endothelial function is regained by valve replacement highlights the importance to maintain regular composition of blood components and thus circulatory endothelial function in aortic valve stenosis, since endothelial dysfunction is a major determinant for poor cardiovascular prognosis.

In view of the growing expansion of indications and the selection of suitable prostheses in increasingly challenging anatomies (e.g. bicuspid valves, valve in valve), knowledge about the impact of disturbed flow conditions on the entire organism is essential and highly relevant.

## Data Availability

All data produced in the present work are contained in the manuscript.

## Data Availability

All data produced in the present work are contained in the manuscript.

## Acknowledgement

We thank Julia Odendahl, Stefanie Becher and Kathrin Paul-Krahé for excellent technical support.

## Abbreviations

Ach: Acetylcholine
AS: aortic valve stenosis
AUC: area under the curve
AVA: aortic valve area
AVC: aortic valve calcification
BMI: body mass index
CKD: chronic kidney disease
CO: cardiac output
DM: diabetes mellitus
dP/dt_max_: maximal rate of rise of left ventricular pressure in systole
dP/dt_min_: minimal rate of rise of left ventricular pressure in systole
EF: ejection fraction
fHb: cell-free hemoglobin
FMD: flow mediated dilation
HLP: hyperlipoproteinemia
Hp: haptoglobin
HR: heart rate
HT: hypertension
IVSd: interventricular septum in diastole
LCC: left coronary cusp
LV: left ventricle
LVEF: left ventricular ejection fraction
LVDP: left ventricular developed pressure
LVIDd: left ventricular inner diameter in diastole
LVOT: left ventricular outflow tract
MSCT: multi-slice computed tomography
NCC: non-coronary cusp
NO_3_^-^: nitrate
NO_2_^-^: nitrite
RBCs: red blood cells
RCC: right coronary cusp
RDW: RBC distribution width
SVR: systemic vascular resistance
TAVR: transcatheter aortic valve replacement

## Graphical abstract

**In AS poststenotic aortic swirling blood flow induces occult hemolysis causing endothelial dysfunction**

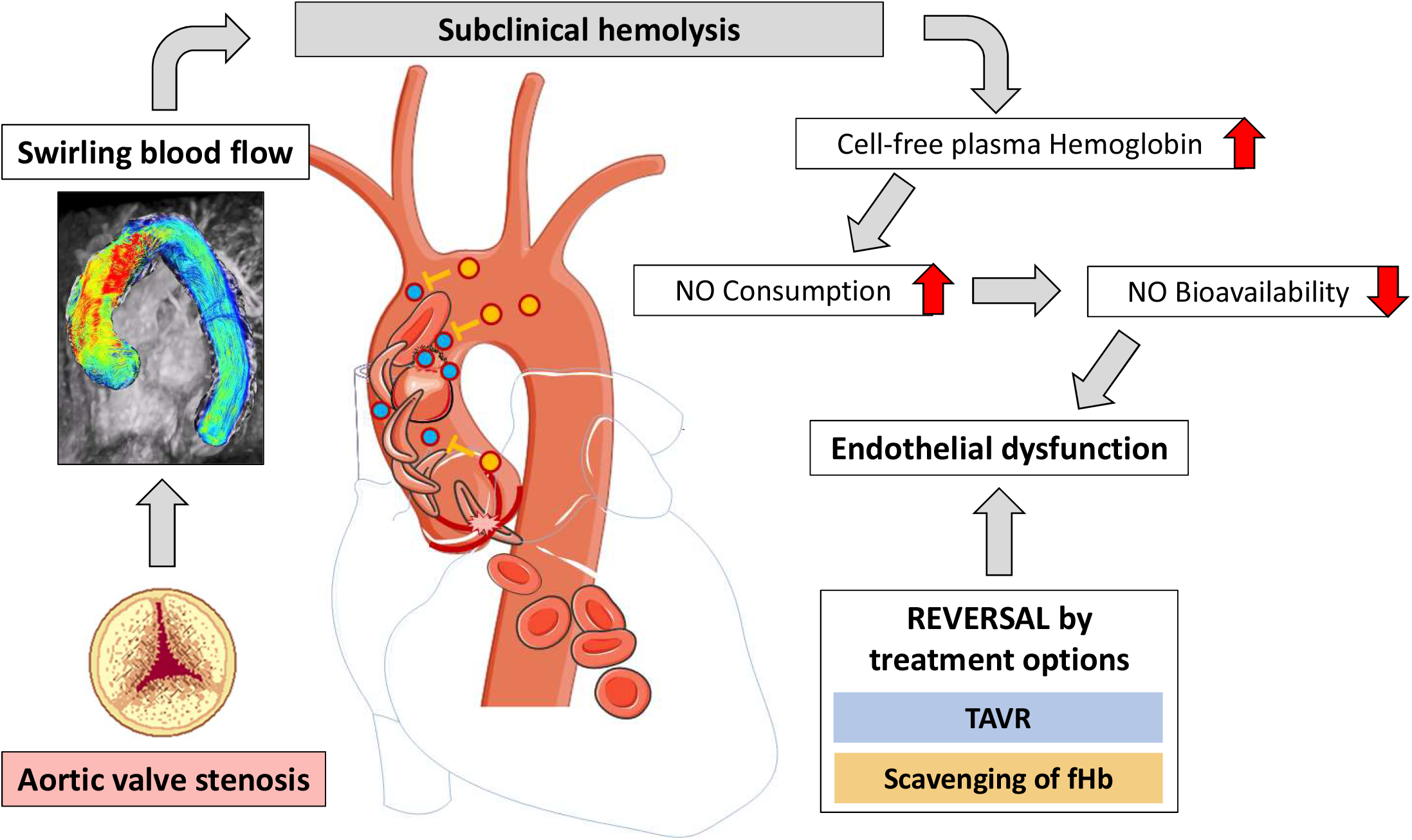

Aortic valve stenosis leads to increased swirling blood flow in the ascending aorta. Increased shear stress acting on RBCs in swirling blood flow above the aortic valve compromise RBC membrane integrity, triggering hemoglobin release. This in turn, reduces NO bioavailability due to increased consumption and NO scavenging by circulating cell-free hemoglobin which translates into endothelial dysfunction. NO bioavailability in plasma is restored by treatment options such as valve replacement (TAVR) or scavenging (yellow dots) of extracellular Hb (fHb, blue dots) which restores endothelial function. (Graphical abstract was partly generated using Servier Medical Art, provided by Servier, licensed under a Creative Commons Attribution 3.0 unported license)

## References

1. Frimat M, Boudhabhay I, Roumenina LT. Hemolysis Derived Products Toxicity and Endothelium: Model of the Second Hit. Toxins. 2019;11(11).

2. Donadee C, Raat NJ, Kanias T, Tejero J, Lee JS, Kelley EE, et al. Nitric oxide scavenging by red blood cell microparticles and cell-free hemoglobin as a mechanism for the red cell storage lesion. Circulation. 2011;124(4):465–76.

3. Arnal JF, Dinh-Xuan AT, Pueyo M, Darblade B, Rami J. Endothelium-derived nitric oxide and vascular physiology and pathology. Cellular and molecular life sciences : CMLS. 1999;55(8-9):1078–87.

4. Vahidkhah K, Cordasco D, Abbasi M, Ge L, Tseng E, Bagchi P, et al. Flow-Induced Damage to Blood Cells in Aortic Valve Stenosis. Annals of biomedical engineering. 2016;44(9):2724–36.

5. Meyer C, Heiss C, Drexhage C, Kehmeier ES, Balzer J, Muhlfeld A, et al. Hemodialysis-induced release of hemoglobin limits nitric oxide bioavailability and impairs vascular function. J Am Coll Cardiol. 2010;55(5):454–9.

6. Honda S, Miyamoto T, Watanabe T, Narumi T, Kadowaki S, Honda Y, et al. A novel mouse model of aortic valve stenosis induced by direct wire injury. Arterioscler Thromb Vasc Biol. 2014;34(2):270–8.

7. Niepmann ST, Steffen E, Zietzer A, Adam M, Nordsiek J, Gyamfi-Poku I, et al. Graded murine wire-induced aortic valve stenosis model mimics human functional and morphological disease phenotype. Clinical research in cardiology : official journal of the German Cardiac Society. 2019.

8. Quast C, Kober F, Becker K, Zweck E, Hoffe J, Jacoby C, et al. Multiparametric MRI identifies subtle adaptations for demarcation of disease transition in murine aortic valve stenosis. Basic Res Cardiol. 2022;117(1):29.

9. Wang X, Tanus-Santos JE, Reiter CD, Dejam A, Shiva S, Smith RD, et al. Biological activity of nitric oxide in the plasmatic compartment. Proceedings of the National Academy of Sciences of the United States of America. 2004;101(31):11477–82.

10. Feelisch M, Rassaf T, Mnaimneh S, Singh N, Bryan NS, Jourd’Heuil D, et al. Concomitant S-, N-, and heme-nitros(yl)ation in biological tissues and fluids: implications for the fate of NO in vivo. Faseb j. 2002;16(13):1775–85.

11. Nogueira RC, Minnion M, Clark AD, Dyson A, Tanus-Santos JE, Feelisch M. On the origin of nitrosylated hemoglobin in COVID-19: Endothelial NO capture or redox conversion of nitrite?: Experimental results and a cautionary note on challenges in translational research. Redox biology. 2022;54:102362.

12. Barker AJ, van Ooij P, Bandi K, Garcia J, Albaghdadi M, McCarthy P, et al. Viscous energy loss in the presence of abnormal aortic flow. Magnetic resonance in medicine. 2014;72(3):620–8.

13. Garcia J, Barker AJ, Markl M. The Role of Imaging of Flow Patterns by 4D Flow MRI in Aortic Stenosis. JACC: Cardiovascular Imaging. 2019;12(2):252–66.

14. Achenbach S, Delgado V, Hausleiter J, Schoenhagen P, Min JK, Leipsic JA. SCCT expert consensus document on computed tomography imaging before transcatheter aortic valve implantation (TAVI)/transcatheter aortic valve replacement (TAVR). J Cardiovasc Comput Tomogr. 2012;6(6):366–80.

15. Clavel MA, Pibarot P, Messika-Zeitoun D, Capoulade R, Malouf J, Aggarval S, et al. Impact of aortic valve calcification, as measured by MDCT, on survival in patients with aortic stenosis: results of an international registry study. J Am Coll Cardiol. 2014;64(12):1202–13.

16. Moscarelli M, Devito F, Fattouch K, Lancellotti P, Ciccone MM, Rizzo P, et al. The effect of surgical versus transcatheter aortic valve replacement on endothelial function. An observational study. International Journal of Surgery. 2019;63:1–7.

17. Gulan U, Luthi B, Holzner M, Liberzon A, Tsinober A, Kinzelbach W. An in vitro investigation of the influence of stenosis severity on the flow in the ascending aorta. Medical engineering & physics. 2014;36(9):1147–55.

18. Kubo T, Okumiya T, Baba Y, Hirota T, Tanioka K, Yamasaki N, et al. Erythrocyte creatine as a marker of intravascular hemolysis due to left ventricular outflow tract obstruction in hypertrophic cardiomyopathy. Journal of cardiology. 2016;67(3):274–8.

19. Sugiura T, Okumiya T, Kubo T, Takeuchi H, Matsumura Y. Evaluation of Intravascular Hemolysis With Erythrocyte Creatine in Patients With Aortic Stenosis. International heart journal. 2016;57(4):430–3.

20. Arashiki N, Takakuwa Y. Maintenance and regulation of asymmetric phospholipid distribution in human erythrocyte membranes: implications for erythrocyte functions. Current opinion in hematology. 2017;24(3):167–72.

21. van Ooij P, Garcia J, Potters WV, Malaisrie SC, Collins JD, Carr JC, et al. Age-related changes in aortic 3D blood flow velocities and wall shear stress: Implications for the identification of altered hemodynamics in patients with aortic valve disease. J Magn Reson Imaging. 2016;43(5):1239–49.

22. van Ooij P, Markl M, Collins JD, Carr JC, Rigsby C, Bonow RO, et al. Aortic Valve Stenosis Alters Expression of Regional Aortic Wall Shear Stress: New Insights From a 4-Dimensional Flow Magnetic Resonance Imaging Study of 571 Subjects. J Am Heart Assoc. 2017;6(9).

23. Farag ES, Vendrik J, van Ooij P, Poortvliet QL, van Kesteren F, Wollersheim LW, et al. Transcatheter aortic valve replacement alters ascending aortic blood flow and wall shear stress patterns: A 4D flow MRI comparison with age-matched, elderly controls. European Radiology. 2019;29(3):1444–51.

24. Trauzeddel RF, Löbe U, Barker AJ, Gelsinger C, Butter C, Markl M, et al. Blood flow characteristics in the ascending aorta after TAVI compared to surgical aortic valve replacement. The international journal of cardiovascular imaging. 2016;32(3):461–7.

25. Peter T, Bissinger R, Lang F. Stimulation of Eryptosis by Caspofungin. Cellular physiology and biochemistry : international journal of experimental cellular physiology, biochemistry, and pharmacology. 2016;39(3):939–49.

26. Dreischer P, Duszenko M, Stein J, Wieder T. Eryptosis: Programmed Death of Nucleus-Free, Iron-Filled Blood Cells. Cells. 2022;11(3).

27. Alfhili MA, Basudan AM, Aljaser FS, Dera A, Alsughayyir J. Bioymifi, a novel mimetic of TNF-related apoptosis-induced ligand (TRAIL), stimulates eryptosis. Medical oncology (Northwood, London, England). 2021;38(12):138.

28. Nikfar M, Razizadeh M, Zhang J, Paul R, Wu ZJ, Liu Y. Prediction of mechanical hemolysis in medical devices via a Lagrangian strain-based multiscale model. Artificial organs. 2020;44(8):E348–e68.

29. Horobin JT, Watanabe N, Hakozaki M, Sabapathy S, Simmonds MJ. Shear-stress mediated nitric oxide production within red blood cells: A dose-response. Clinical hemorheology and microcirculation. 2019;71(2):203–14.

30. Faghih MM, Sharp MK. Modeling and prediction of flow-induced hemolysis: a review. Biomechanics and modeling in mechanobiology. 2019;18(4):845–81.

31. Horn P, Stern D, Veulemans V, Heiss C, Zeus T, Merx MW, et al. Improved endothelial function and decreased levels of endothelium-derived microparticles after transcatheter aortic valve implantation. EuroIntervention : journal of EuroPCR in collaboration with the Working Group on Interventional Cardiology of the European Society of Cardiology. 2015;10(12):1456–63.

32. Güz G, Demirgan S. Lower brachial artery flow-mediated dilation is associated with a worse prognosis and more lung parenchymal involvement in Covid-19: Prospective observational study. Medicine. 2022;101(33):e30001.

33. Melo R, f AS, Ferreira AF, Moreira S, Moreira R, R JC, et al. Endothelial Function and Vascular Properties in Severe Aortic Stenosis Before and After Aortic Valve Replacement Surgery. Revista portuguesa de cirurgia cardio-toracica e vascular : orgao oficial da Sociedade Portuguesa de Cirurgia Cardio-Toracica e Vascular. 2017;24(3-4):153.

34. Jung C, Lichtenauer M, Figulla HR, Wernly B, Goebel B, Foerster M, et al. Microparticles in patients undergoing transcatheter aortic valve implantation (TAVI). Heart and vessels. 2017;32(4):458–66.

35. Morris ST, Jardine AG. The vascular endothelium in chronic renal failure. Journal of nephrology. 2000;13(2):96–105.

36. Radenkovic M, Stojanovic M, Prostran M. Endothelial Dysfunction in Renal Failure: Current Update. Current medicinal chemistry. 2016;23(19):2047–54.

37. Ter Maaten JM, Damman K, Verhaar MC, Paulus WJ, Duncker DJ, Cheng C, et al. Connecting heart failure with preserved ejection fraction and renal dysfunction: the role of endothelial dysfunction and inflammation. European journal of heart failure. 2016;18(6):588–98.

38. Zuchi C, Tritto I, Carluccio E, Mattei C, Cattadori G, Ambrosio G. Role of endothelial dysfunction in heart failure. Heart failure reviews. 2020;25(1):21–30.

39. Collins P, Burman J, Chung HI, Fox K. Hemoglobin inhibits endothelium-dependent relaxation to acetylcholine in human coronary arteries in vivo. Circulation. 1993;87(1):80–5.

40. Deuel JW, Vallelian F, Schaer CA, Puglia M, Buehler PW, Schaer DJ. Different target specificities of haptoglobin and hemopexin define a sequential protection system against vascular hemoglobin toxicity. Free radical biology & medicine. 2015;89:931–43.

41. Schaer CA, Deuel JW, Bittermann AG, Rubio IG, Schoedon G, Spahn DR, et al. Mechanisms of haptoglobin protection against hemoglobin peroxidation triggered endothelial damage. Cell death and differentiation. 2013;20(11):1569–79.

42. Boulanger CM, Amabile N, Guerin AP, Pannier B, Leroyer AS, Mallat CN, et al. In vivo shear stress determines circulating levels of endothelial microparticles in end-stage renal disease. Hypertension. 2007;49(4):902–8.

43. Camus SM, De Moraes JA, Bonnin P, Abbyad P, Le Jeune S, Lionnet F, et al. Circulating cell membrane microparticles transfer heme to endothelial cells and trigger vasoocclusions in sickle cell disease. Blood. 2015;125(24):3805–14.

44. Marchini JF, Miyakawa AA, Tarasoutchi F, Krieger JE, Lemos P, Croce K. Endothelial, platelet, and macrophage microparticle levels do not change acutely following transcatheter aortic valve replacement. Journal of negative results in biomedicine. 2016;15:7.

